# Women’s Awareness of Obstetric Fistula and its Associated Factors Among Reproductive-Age Women in Ethiopia: A Multilevel Analysis Based on National Survey Data

**DOI:** 10.1101/2021.04.29.21256293

**Authors:** Wallelign Alemnew, Bezawit Mulat, Kegnie Shitu

## Abstract

**Objective:** This study aimed to determine the magnitude and associated factors of women’s awareness of obstetric fistula (OF).

**Methods:** This community based crossectional study was conducted among 15,683 reproductive-aged women. A multi-level multivariable logistic regression analysis employed. A 95% CI and p-value < 0.05 were used to declare statistical significance.

**Result:** The magnitude of women’s awareness of OF was 38% [95%CI: 0.37, 0.39]. Individual level variables such as being in the age group 20-25 [AOR=1.17; 95%CI:1.02, 1.35], 26-30 [AOR=1.50; 95%CI: 1.27, 1.76] and >30 [AOR=1.76; 95%CI: 1.50, 2.07], being Muslim religion follower [AOR = 0.83; 95%CI: 0.73, 0.94], attending primary education [AOR=1.70; 95%CI: 1.53, 1.89], attending secondary education [AOR=3.43; 95%CI: 2.95, 3.99] and attending higher education [AOR=5.88; 95%CI: 4.66, 7.42], history of pregnancy termination [AOR=1.31; 95%CI: 1.13, 1.51], media exposure at least once a week [AOR= 1.33; 95%CI: 1.2, 1.49], internet use [AOR= 2.25; 95%CI: 1.84, 2.75], medium house hold wealth [AOR=1.17; 95%CI: 1.02, 1.34], rich house hold wealth [AOR=1.50; 95%CI:1.31, 1.72] and community level factors including high community media exposure [AOR = 1.30; 95%CI: 1.05, 1.61], high community ANC rate [AOR=1.66; 95%CI: 1.37, 2.02] and low health facility distance problem [AOR=1.49; 95%CI: 1.23, 1.81] were significantly associated with awareness of OF.

**Conclusion:** The magnitude of women’s awareness of OF was very low in Ethiopia. Maternal age, religion, being educated, media exposure, being from a wealthier household, low health facility distance problem, and high community ANC rate were significantly and positively associated factors with awareness of OF.

## Background

Obstetric fistula is a life-altering birth associated injury [1] that creates an abnormal connection between the genital tract and the urinary tract (urogenital fistula) or the gastrointestinal tract (most commonly, rectovaginal fistula) that occur as the result of obstetric trauma, typically from prolonged obstructed labor [2, 3]. It is a public health concern for women and their communities within developing nations, particularly in Africa and Southeast Asia [4] and it is found to be one of the most visible indicators of maternal morbidity [5]. The World Health Organization estimates there are 130,000 new cases of obstetric fistula each year, calculated from an assumption that fistula is likely to occur in 2 percent of the 6.5 million cases of obstructed labor that occur in developing countries [2, 6]. Of these, around 33,000 women are found in Sub-Saharan Africa, and it affects about 1.62 women per 1000 reproductive age in Ethiopia [7, 8]. Currently around 36,000 and 39,000 women living with obstetric fistula and that every year there are between 3,300 and 3,750 new cases of obstetric fistula in Ethiopia according to the problem analysis report [9].

Pieces of evidence show that the risk of the vaginal fistula is common in settings with a lack of inadequate emergency obstetric care, healthcare workforce shortages, and poor investment in maternity services [6, 10]. Further, sociocultural issues such as early marriage, harmful cultural practices like female genital mutilation, home-based childbirth, poor policy implementation of female education, and misconceptions about childbirth practices are other contributing factors for the high burden of obstetric fistula in Sub-Saharan Africa [11, 12]. Adequate information and awareness of the risk factors, causes, and treatment options for vaginal fistula may help women to take appropriate steps to prevent vaginal fistula. Generally, different researches reported that still there is a low level of awareness about obstetric fistula in Africa, especially in Ethiopia. The magnitude of awareness was found to be 40.8% and 39.5% in Bench sheka zone, south, Ethiopia, and awi zone Amhara region, Ethiopia respectively [7, 9].

Despite the high burden of obstetric fistula in Sub-Saharan Africa, studies addressing the awareness of obstetric fistula among women are limited, particularly in Ethiopia. Therefore, this study aims to determine the magnitude of obstetric fistula awareness and factors that could contribute to the awareness of obstetric fistula among women of reproductive age in Ethiopia. Furthermore, findings from this analysis will assist policy-makers and public health programmers to understand the magnitude of obstetric fistula awareness and the contributory factors.

## Methods

### Data source, population, and sampling procedure

The present study was based on the most recent Ethiopian Demographic and Health Survey (EDHS) data. The survey was conducted from January 18, 2016, to June 27, 2016. For this survey, a complete list of 84, 915 Enumeration Areas (EAs) from the Ethiopian Population and Housing Census was used as a sampling frame. A stratified two-stage cluster sampling technique was employed. In the first stage, 645 EAs were selected. In the second stage, an average of 28 households was selected per cluster/EA. The data is freely available in public and we accessed the dataset used for the present study after we registered and received an authentication letter from the Demographic and Health Survey (DHS) program at The DHS Program - Ethiopia: Standard DHS, 2016 Dataset. For this study, a total weighted sample of 15, 683 reproductive-age women were included. Detailed information about the sampling strategy, questioner, or other important information is available in the 2016 EDHS report.

### Variables of the Study

The outcome variable of the present study was women’s awareness of obstetric fistula. The variable was dichotomized into **1** = “ever heard of fistula” and **0** = “never heard of fistula”. The independent variables were further classified into individual level (level 1) variables and community level (level 2) variables. Individual-level variables included age, marital status, religion, educational status, media exposure, internet access, family wealth index, sexual activity, birth history, pregnancy termination, current working status, and current pregnancy status. Whereas, community variables involved variables directly taken with no aggregation (residence and contextual region), and variables obtained by aggregating individual values into their respected community (community poverty, community female education, community media exposure, community antenatal care service utilization rate, and community health facility distance). Since the aggregate values of each variable didn’t follow a normal distribution curve, we categorized the aggregate values of a cluster into groups based on median values.

### Operational definitions

#### Household wealth quintile

The wealth index classifications were done in quintiles: poorest; poor; middle; rich; richest. These were computed using principal component analyses (PCA). This variable was further categorized into three categories (Poor, Medium, and Rich) by merging the lower two (poorest and poor) quantiles and the upper two (richest and rich) quantiles.

#### Media Exposure

This variable was computed from the frequency of exposure to the two commonest mass media routes (radio and television). In this study exposure to magazines/newspapers was excluded because little (<5%) women were exposed to this channel. The variable was categorized into three categories: no exposure to either media route; exposure to either one of the two routes; and exposure to both within a week.

#### Contextual region

For this survey regions were categorized into three categories (agrarian, pastoral, and metropolitan) that may have a strong relationship to health information seeking and awareness of obstetric fistula. The Tigray, Amhara, Oromia, SNNP, Gambella, and Beneshangul Gumuz were recorded as agrarian. The Somali and Afar regions were merged to form the pastoralist region and the city administrations-Addis Ababa, Dire Dawa, and Harar were combined as metropolitan [1].

#### Community female education

This is the aggregate value of the educational levels of women based on the average of proportions of educational levels in the community. It was defined as low if the proportion of women with secondary education & above in the community was 0 – 12.4 % and high if the proportion was 12.5 –100 %.

#### Community media exposure

This variable was derived from the individual responses for exposure to radio or television. It was defined as low if the proportion of women exposed to media in the community was 0–18.7 % and high if the proportion was 18.8–100 %.

#### Community ANC utilization rate

This variable is also derived from the individual values for ANC utilization. It was defined as low if the proportion of women who attended at least one ANC visit in the community was 0 – 81.3 % and high if the proportion was between 81.4 –100 %.

#### Community poverty

With the same procedure, this variable is also derived from an individual household’s wealth index. It was defined as high if the proportion of women from the two lowest wealth quintiles in a given community was 25.9–100 % and low if the proportion was 0–26 %.

#### Community distance to the health facility

The variable was aggregated from individual perceived distance to a health facility is a big problem. It was as categorized as low if the proportion of women who perceived health facility distance as a big problem in the community was 0–42.2% and categorized as high if the proportion was between 42.2% and 100%.

### Data processing and Analysis

Data were extracted from individual records (IR) files and further coding and transformations were done using statistical software, STATA version 14. The weighted samples were used for analysis to adjust for unequal probability of selection and non-response in the original survey. Since the EDHS employed multi-stage stratified cluster sampling techniques, the data have a hierarchical structure. Therefore, a multivariable multilevel binary logistic regression model was used to estimate the fixed and random effects of the factors associated with awareness of fistula. Four models were constructed. The first model also called an empty model which was fitted without any explanatory variables. This model is important for understanding the community variations, and we used it as a reference to estimate how much community factors were able to explain the observed variations in the awareness of fistula. Moreover, this model was used to justify the use of a multilevel statistical framework as it is a litmus paper on whether multilevel or the traditional logistic regression should be used. It was assessed using the Likelihood Ratio test (LR), Median Odds Ratio (MOR), Intra-class Correlation Coefficient (ICC), and Proportional Change of Variance (PCV). The second model contained only individual-level factors. The third contained only community-level factors. Whereas, the final (fourth) model containing both individual and community-level factors. Moreover, the model comparison was done using model deviance, a model with the lowest deviance selected for reporting and interpretation results.

## Result

### Individual-level factors

A total of 15683 reproductive-age women were included in this study. The median age of the participants was 27 with an inter-quartile range of 15. Nearly half (47.8%) of the participants didn’t attend any formal education and more than half (63.8%) of them were married. Only one-third (33.3%) of them were employed during the survey. Regarding the reproductive history, about 77% (12033), 67.5 (10587), and 7.2% (1135) of the participants were ever had sex, ever had a child, and currently pregnant respectively. Moreover, only 18.5% (2,896), and 4.4% (693) of women had been exposed to either television or radio at least once a week and had ever used the internet respectively (Table 1).

**Table 1:** Sociodemographic and sexual characteristics of the reproductive age women in Ethiopia (n = 15, 683)

| Variables | Frequency | Proportion |
| --- | --- | --- |
| Age |  |  |
| 15-19 | 3, 381 | 21.6 |
| 20-25 | 3, 655 | 23.3 |
| 26-30 | 2, 882 | 18.4 |
| >=31 | 5, 765 | 36.7 |
| Religion |  |  |
| Cristian | 10, 580 | 67.4 |
| Muslim | 4, 893 | 31.1 |
| Others | 205 | 1.3 |
| Highest educational level |  |  |
| No-formal education | 7498 | 47.8 |
| Primary | 5490 | 35 |
| Secondary | 1818 | 11.6 |
| Higher | 877 | 5.6 |
| Marital status |  |  |
| Married | 10014 | 63.8 |
| Unmarried | 5669 | 36.2 |
| Ever had a child |  |  |
| No | 5096 | 32.5 |
| Yes | 10587 | 67.5 |
| Sexual experience |  |  |
| Never had sex | 3650 | 23.3 |
| Ever had sex | 12033 | 76.7 |
| Currently pregnant |  |  |
| No or unsure | 14548 | 92.8 |
| Yes | 1135 | 7.2 |
| Ever had a terminated pregnancy |  |  |
| No | 14447 | 92.1 |
| Yes | 1236 | 7.9 |
| Mass media exposure (TV/Radio) |  |  |
| Both less than a week or not at all | 11,702 | 74.6 |
| Either at least once a week | 2,896 | 18.5 |
| Both at least once a week | 1,085 | 6.9 |
| Use of Internet |  |  |
| Never | 14,904 | 95.0 |
| Ever | 693 | 4.4 |
| Currently working |  |  |
| No | 10,463 | 66.7 |
| Yes | 5,220 | 33.3 |
| Household wealth Index |  |  |
| Poor | 8,420 | 53.7 |
| Medium | 3,100 | 19.8 |
| Rich | 4,164 | 26.5 |

### Community-level variables

The majority (77.8%) of the participants were from rural residency. About 50.8% (7,714), 60.3% (9,452), 58.0% (9,089), and 62.8% (9,848) were from a community where there is high poverty low female education, low media exposure and low ANC utilization rate respectively (Table 2).

**Table 2:** Summary result of community-level factors of women’s awareness of obstetric fistula in Ethiopia (n=15, 683)

fistula in Ethiopia (n=15, 683)
| Variables | Frequency | Proportion |
| --- | --- | --- |
| Residence |  |  |
| Urban | 3,476 | 22.2 |
| Rural | 12,207 | 77.8 |
| Contextual Region |  |  |
| Agrarian | 13,870 | 88.5 |
| Pastoral | 792 | 5.0 |
| Metropolitan | 1,021 | 6.5 |
| Community poverty |  |  |
| Low | 7,969 | 49.2 |
| High | 7,714 | 50.8 |
| Community female education |  |  |
| Low | 9,452 | 60.3 |
| High | 6,231 | 39.7 |
| Community media exposure |  |  |
| Low | 9,089 | 58.0 |
| High | 6,594 | 42.0 |
| Community ANC utilization rate |  |  |
| Low | 9,848 | 62.8 |
| High | 5,835 | 37.2 |
| Distance to Health facility is a big problem |  |  |
| Low | 6,656 | 42.4 |
| High | 9,027 | 57.6 |

### Awareness of obstetric fistula

The overall prevalence of awareness of fistula among reproductive-age women was 38% (95% CI: 37%-39%).

### Random effect and Model comparison

As is depicted in table 4 the null model revealed that awareness of fistula was significantly varied across communities/clusters. This was assessed by the ICC value in the null model by which 36.0% of the total variation in awareness of fistula was due to the difference between clusters. Moreover, the MOR in the null model, which was 3.63 (95% CI: (3.32, 3.97)), indicated that there was a significant variation between clusters. This shows if a woman moves to another area with a higher probability of awareness of fistula, her risk of having awareness of fistula will increase 3.63 times. The final model that included both the individual and community level factors explained the greatest variance (52 %) in fistula awareness as it was indicated by PCV. All of the three parameters revealed that the effect of clustering was significant. Indeed, based on model fit comparison indices, the final model was found to be the best-fitted model (had the lowest deviance) (Table 3).

**Table 3:**
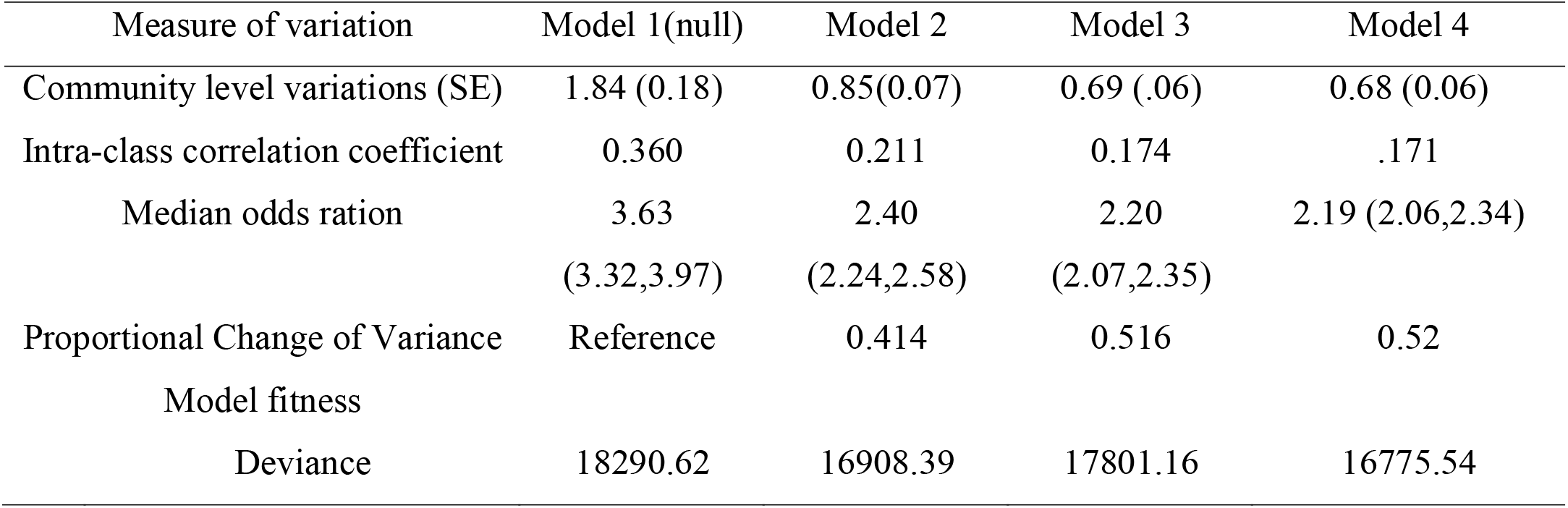
Random effect model and model fitness for factors associated with women’s awareness of obstetric fistula in Ethiopia (n=15, 683)

**Table 4:**
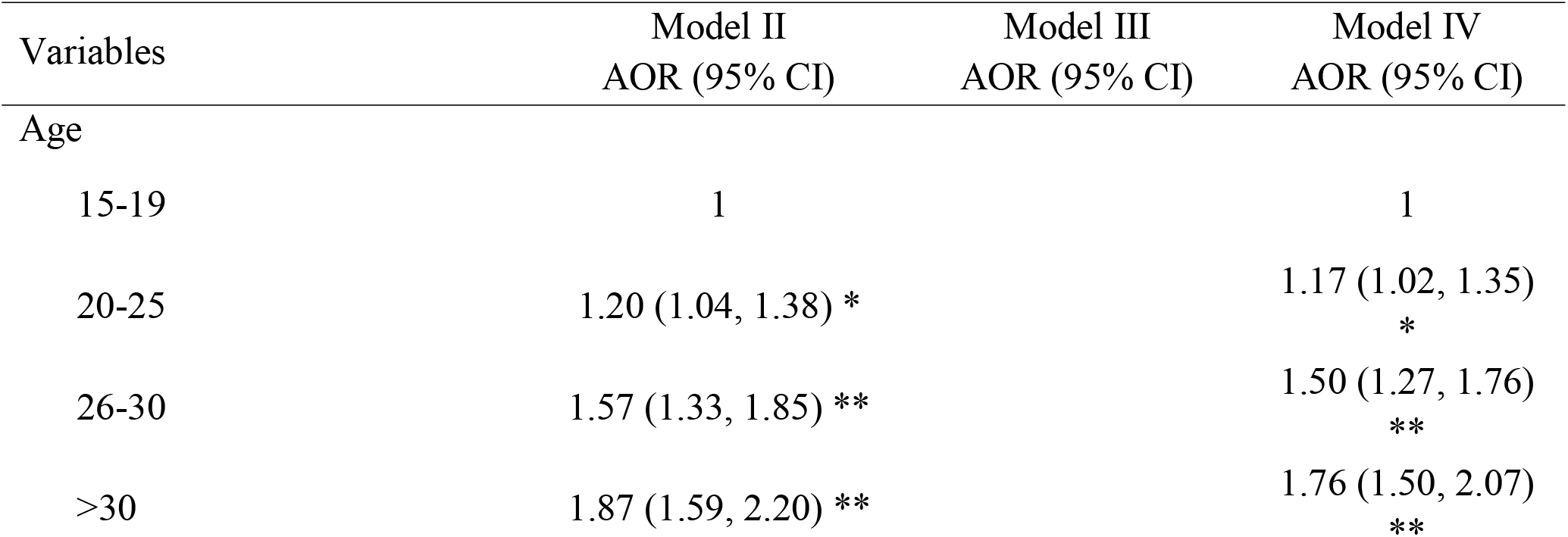
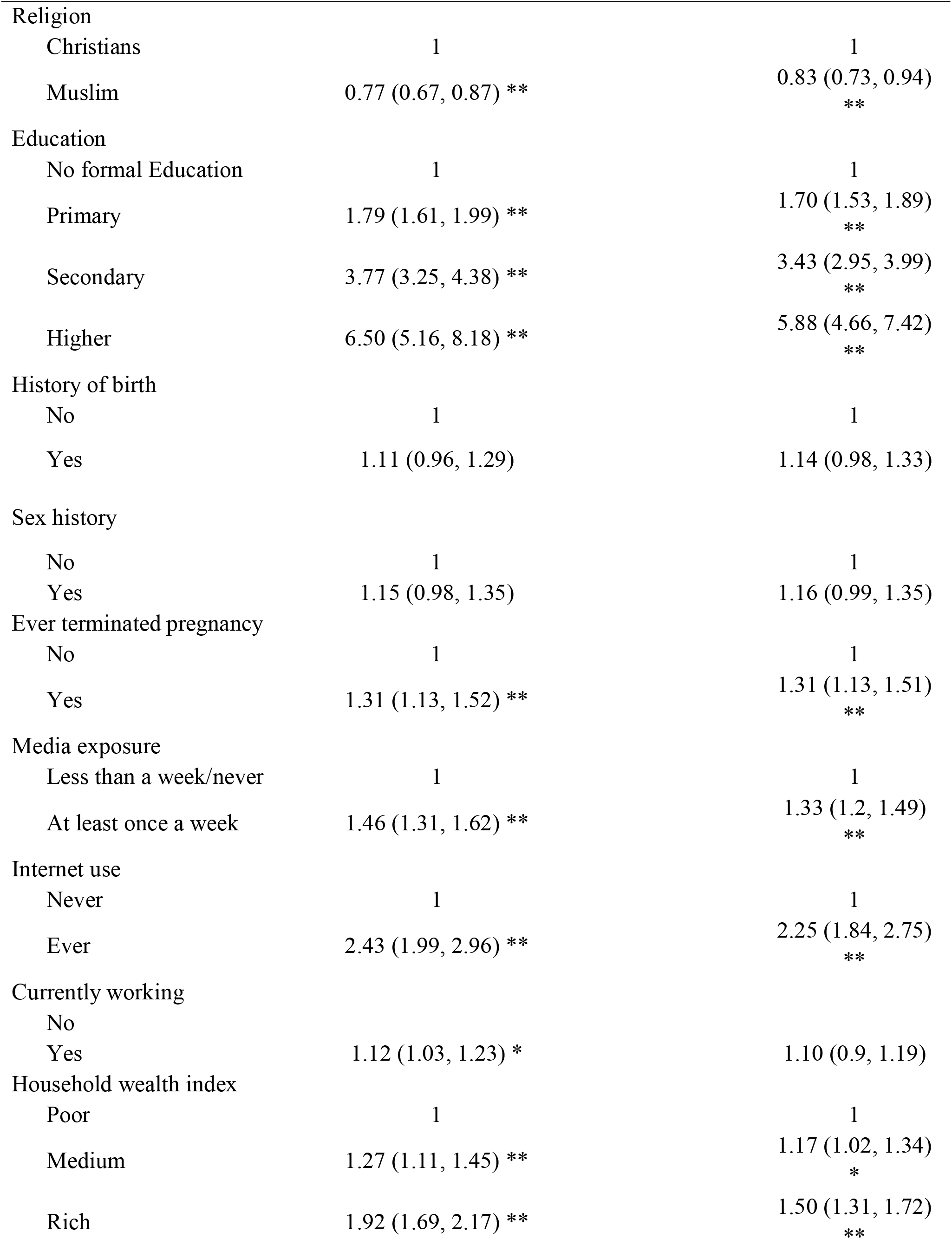
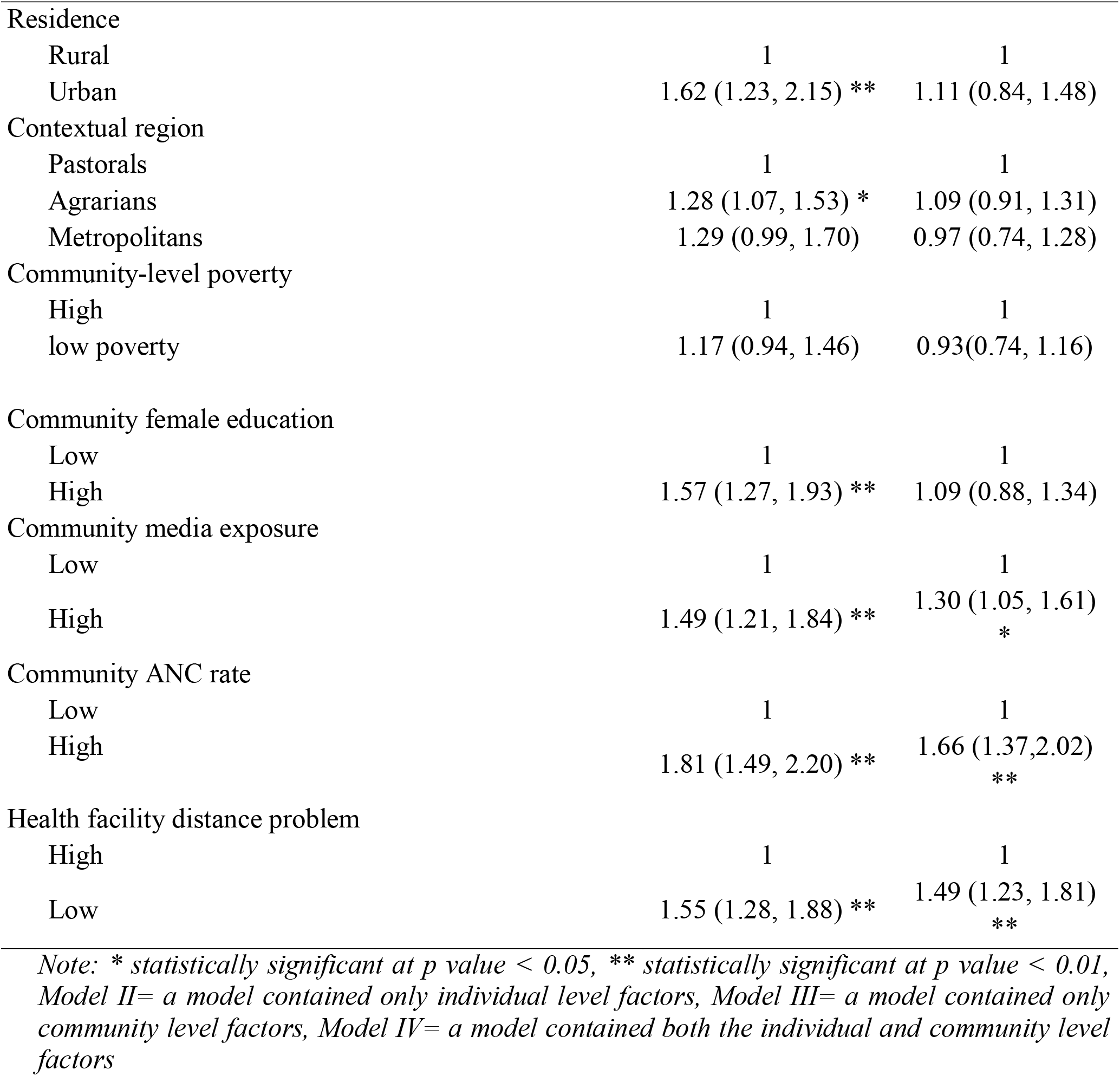
A multilevel logistic regression analysis of factors associated with women’s awareness of obstetric fistula among reproductive age women in Ethiopia (n=15, 683)

### Individual and community level factors associated with women’s awareness of obstetric fistula

The multivariable multilevel logistic regression analysis showed that age, religion, education, ever pregnancy termination, media exposure, internet use, household wealth index, community media exposure, community health facility distance problem, and community ANC utilization rate were significantly associated with women’s awareness of obstetric fistula at p-value <0.05.

The odds of having awareness of fistula was higher among women with the age of 20-25 years old (AOR =1.17, 95% CI: 1.02, 1.35)), 26-30 years old (AOR= 1.5, 95% CI: (1.27, 1.76)) and > 30 years old (AOR= 1.76, 95% CI: 1.5, 2.07) as compared to women with the age of 15-19 years old. Women of Islamic religion followers had 17% lower odds of having fistula awareness (AOR= 0.83, 95% CI: (0.73, 0.94) than Christianity followers. Women with primary (AOR= 1.7, 95% CI: (1.53, 189)), secondary (AOR= 3.43; 95% CI: (2.95, 3.99)), and tertiary education (AOR= 5.88; 95% CI: (4.66, 7.42)) had higher odds of having awareness of fistula than women with no formal education respectively. Moreover, the odds of having fistula awareness were higher among women who had ever terminated pregnancy than their counterparts (AOR=1.31, 95% CI: (1.13, 1.51)). Women who were exposed to radio and /or TV at least once a week were more likely to have awareness of fistula than those who were exposed less than a week/ not at all (AOR = 1.33 95% CI: (1.2, 1.49)). Moreover, the odds of having awareness of fistula were 2.3 times higher among women who had ever used the internet (AOR= 2.25, 95% CI: (1.84, 2.75)). Regarding Woman’s family wealth index, a woman from a family with the highest two quantiles (rich) and third quantile (medium) was more likely to have awareness of fistula as compared to a woman from a family with the lowest two quantiles (AOR=1.17, 95% CI: (1.02, 1.34)) and (AOR= 1.50, 95% CI: (1.31, 1.72)) respectively. The odds of having fistula awareness were 1.3 (AOR=1.30 95%CI: (1.05, 1.61)), 1.66 (AOR=1.66, 95% CI: (1.37, 2.02)) and 1.49 (AOR=1.49, 95% CI: (1.23, 1.81) times higher among a woman from a community with high media exposure, high ANC utilization, and high community health distance problem respectively (Table 4).

## Discussion

The study aimed to assess the magnitude and associated factors of awareness of fistula in Ethiopia using EDHS 2016 data. According to the findings of this study, the estimated magnitude of fistula awareness in Ethiopia was 38% which is consistent with studies done in Bench sheka zone, south, Ethiopia, and Awi zone Amhara region Ethiopia 40.8% and 39.5% respectively [7, 9]. The possible justification for the similarity might be due to the same socio-demographic characteristic of the respondents. Whereas the finding is lower than studies conducted in different countries like Nigeria (57.8%), Ghana (45.8%), Nepal (72.99%), and Dabat, northwest, Ethiopia (54.7%)[8, 13-16]. The difference might be due to the study period, study population, study design, and socio-demographic characteristics. In addition to these, the above studies were based on small sample size or they are mostly facility and community-based cross-sectional studies.

In multivariable multilevel logistic regression analysis being in the age group 20-25, 26-30 years and, being Muslim religion follower, attending primary education, attending secondary education and attending higher education, history of pregnancy termination, media exposure at least once a week, internet use, medium household wealth, rich households wealth, community ANC and low health facility distance were significantly associated with fistula awareness in Ethiopia.

Older women had high odds of fistula awareness compared to young age women; this is might be due to that older women have had different or multiple exposures to media, different health education forums and they have had better opportunities to access/engage in health care services including counseling services [11]. Attending formal education has had higher odds than not attending formal education for fistula awareness. The finding agrees with studies done in the Awi zone of North West Ethiopia, Bench sheka zone, south, Ethiopia Burkina Faso, and Northern Ghana [7, 9, 11]. The possible explanation for these might be those attending formal education have greater opportunities to get information, asking, and getting health services than those who hadn’t attended formal education.

Besides, our study reveals that previous experience or exposure to pregnancy termination had statistical significance with the level of fistula awareness. Women who got pregnancy termination history or experience were 1.31 times more likely to have a better awareness than those who hadn’t get pregnancy termination experiences. The possible justification may be getting or seeking health care services may create an indirect opportunity for counseling and health education; these may increase the level of awareness or their knowledge towards fistula.

In this study, community ANC rate has a significant association with awareness on fistula, those who have high community ANC rate have high odds compared to low community ANC rate. The finding is more supported with a study conducted in the Amhara region, Delanta woreda and Asela, Oromia region [17, 18] this implies that more awareness regarding fistula might be linked to ANC coverage increment where women can be counseled about maternal health issues.

Other important factors which have a significant association with fistula awareness were wealth quin-tiles, and access to information dissemination platforms (mobile and internet utilization) in this study, which had also been previously reported in studies [19-22] conducted in different areas.

In this study, there was a significant association between women’s awareness about fistula and distance to the nearby health facility. For women who perceived health, facility distance as a low problem in the community, their awareness is 1.55 more likely to be good than those women who perceived health facility distance as a high problem. The finding is supported by the studies in Ethiopia and China respectively [23, 24] that identify the distance to the health care facility was linked with poor knowledge related to fistula and poor health-seeking behavior. This might be due to as the health facility becomes near; women are willing to visit the health care facilities and have got a chance to attend health-related information and education.

## Conclusion and recommendations

The magnitude of awareness towards fistula was very low in Ethiopia. Maternal age, religion, being educated, media exposure, from high wealth family, low distance of health facility problem, and high community ANC rate are factors that were significantly and positively associated with women’s awareness towards fistula. Thus, community awareness creation actions are mandatory to offer accurate information on the risk factors, causes, and treatment options of obstetric fistula.

## Data Availability

All the data sets are publicly available at www. dhsmeasure.com upon reasonable request.

## List of abbreviations

ANC: Antenatal care
BCC: Behavioral Change Communication
EDHS: Ethiopian Demographic and Health Survey
AOR: Adjusted Odds Ratio
ICC: Intra Class Correlation
MOR: Median Odd Ratio PCV Proportional Change in Variance

## Ethics approval

We registered our research project at the Demographic and Health Survey by submitting a concept not concerning the topic of interest. After evaluation of our concept note the International Review Board of DHS program data archivists gave us a permission letter to download use the dataset for this registered research project.

## Consent for publication

Not applicable.

## Availability of data and materials

The dataset used for the analysis the present study is freely available at: https://dhsprogram.com

## Competing interests

The authors declare that they have no competing interests

## Funding

There was no fund/grant received for this research project

## Authors’ contributions

WA, BM and KS were involved in inception, methods and acquisition of the data. KS did data extraction and formal analysis. WA and BM did the interpretation of results and writing the draft manuscript. WA, BM and KS were involved in project administration and revising the final manuscript. All authors read and approved the final manuscript.

## Acknowledgments

We would like to acknowledge the MEASURE DHS program for permitting us to obtain and use dataset used for this study.

## Notes

### Competing Interest Statement

The authors have declared no competing interest.

### Funding Statement

There was no received fund for this study.

### Author Declarations

Since this study was based on the secondary data, permission from the data owner(DHSMEASURE) was obtained after providing reasonable justification of the study

